# SARS-CoV-2 Seroprevalence and Drug Use in Trauma Patients from Six Sites in the United States

**DOI:** 10.1101/2021.08.10.21261849

**Authors:** Tran B. Ngo, Maria Karkanitsa, Kenneth M. Adusei, Lindsey A. Graham, Emily E. Ricotta, Jenna R. Darrah, Richard D. Blomberg, Jacquelyn Spathies, Kyle J. Pauly, Carleen Klumpp-Thomas, Jameson Travers, Jennifer Mehalko, Matthew Drew, Matthew D Hall, Matthew J Memoli, Dominic Esposito, Rosemary A. Kozar, Christopher Griggs, Kyle W. Cunningham, Carl I. Schulman, Marie Crandall, Mark Neavyn, Jon D. Dorfman, Jeffrey T. Lai, Jennifer M. Whitehill, Kavita M. Babu, Nicholas M. Mohr, Jon Van Heukelom, James C. Fell, Whit Rooke, Heather Kalish, F. Dennis Thomas, Kaitlyn Sadtler

## Abstract

In comparison to the general patient population, trauma patients show higher level detections of bloodborne infectious diseases, such as Hepatitis and Human Immunodeficiency Virus. In comparison to bloodborne pathogens, the prevalence of respiratory infections such as SARS-CoV-2 and how that relates with other variables, such as drug usage and trauma type, is currently unknown in trauma populations. Here, we evaluated SARS-CoV-2 seropositivity and antibody isotype profile in 2,542 trauma patients from six Level-1 trauma centers between April and October of 2020 during the first wave of the COVID-19 pandemic. We found that the seroprevalence in trauma victims 18-44 years old (9.79%, 95% confidence interval/CI: 8.33–11.47) was much higher in comparison to older patients (45-69 years old: 6.03%, 4.59-5.88; 70+ years old: 4.33%, 2.54 – 7.20). Black/African American (9.54%, 7.77 – 11.65) and Hispanic/Latino patients (14.95%, 11.80 – 18.75) also had higher seroprevalence in comparison, respectively, to White (5.72%, 4.62–7.05) and Non-Latino patients (6.55%, 5.57 – 7.69). More than half (55.54%) of those tested for drug toxicology had at least one drug present in their system. Those that tested positive for narcotics or sedatives had a significant negative correlation with seropositivity, while those on anti-depressants trended positive. These findings represent an important consideration for both the patients and first responders that treat trauma patients facing potential risk of respiratory infectious diseases like SARS-CoV-2.

## INTRODUCTION

The Coronavirus Disease 2019 (COVID-19) pandemic has been a daunting medical challenge for scientists, clinicians, and healthcare professionals due to the ability of the SARS-CoV-2 virus to spread quickly and, frequently, undetected. Currently, there are over 200 million confirmed cases of COVID-19 globally, with the United States accounting for almost 18 % of these cases^1^. The U.S. prevalence and disparities of SARS-CoV-2 infection have been documented in different demographics and regional areas^2-7^. However, this statistic undercounts pre-symptomatic and asymptomatic patients, both of whom can transmit SARS-CoV-2^8^; hence, the number of people spreading SARS-CoV-2 at any given time is difficult to determine.

Furthermore, there is limited information regarding the prevalence of COVID-19 in patients admitted to hospitals due to trauma. Previous studies indicate that trauma victims have a higher prevalence of certain viral infections, such as Human Immunodeficiency Virus (HIV). In 2018, researchers showed that 1.1% of 1217 individuals in a trauma cohort tested positive for HIV, which was more than three times the national prevalence estimated by the Centers for Disease Control and Prevention (CDC) of the U.S. general population (0.37% or 1.2 million HIV positive cases)^9, 10^. Other viral infection prevalence is also higher in trauma patients than the national average. In a study analyzing positivity of bloodborne viruses Hepatitis B/C and HIV, 75% of patients who tested positive were undiagnosed for these diseases prior to enrollment^11^. Injury severity, another pre-hospital factor, has been shown to be an independent predictor of ventilator-associated pneumonia causing complications in trauma population^12^. Overall, trauma patients require direct and intensive care from many health care providers including the first responders (e.g., emergency medical services/EMS, law enforcement), primary trauma team (e.g., treating medical staff in trauma centers), and specialists (e.g. respiratory, physical, and occupational therapy)^13^.

With a high community transmission rate of SARS-CoV-2 virus along with many variant lineages of concern, first responders and health care workers could be facing a much higher risk of exposure to viral infection than previously expected when treating trauma patients. As reported, COVID-19 related fatality risks were the single highest cause of officer line-of-duty deaths^14, 15^. EMS providers, who have been operating on the far-forward front lines of the pandemic in 2020, had more cases of severe COVID-19 than firefighters (1.2% versus 0.19% respectively)^16^. This risk could be exacerbated by the elevated ability of SARS-CoV-2 to be transmitted by asymptomatic patients. Byambasuren et al. reported a 17% asymptomatic SARS-CoV-2 infection rate of total confirmed SARS-CoV-2 infected patients in a meta-analysis of data from seven countries^17^. Additionally, previous research found that in the summer of 2020, there were approximately 4.8 undiagnosed SARS-CoV-2 infections for every reported case, totaling almost 17 million undiagnosed infections^18^. Since there are a high prevalence of viral infections in trauma population and numerous asymptomatic SARS-CoV-2 cases in the general population, more information is needed to determine if first responders and trauma center staff could be at increased risk. Therefore, knowing the prevalence of COVID-19 among trauma patients would allow first responders and healthcare staff to better assess their risk of SARS-CoV-2 infection to create effective measures to mitigate the risk, along with considerations for their patients.

This study assesses the SARS-CoV-2 seropositivity of 2,542 de-identified serum samples from trauma patients using a standardized enzyme-linked immunosorbent assay (ELISA) protocol that was previously developed for the national serosurvey, conducted May 10^th^ and July 31^st^, 2020^18, 19^. The serosurvey ELISA protocol identified IgG, IgM, and IgA antibodies for the SARS-CoV-2 spike protein and its receptor binding domain (RBD). This assay can assess SARS-CoV-2 seropositivity objectively using either IgG or IgM detected levels – for both spike and RBD expression – based on a threshold determined by pre-pandemic control samples. The goal of this study was to evaluate SARS-CoV-2 seroprevalence in trauma patients to offer insightful information on the association between SARS-CoV-2 infection and trauma, which has not been previously reported. This serological study provides an in-depth assessment of SARS-CoV-2 seropositivity in trauma patients as well as detects different anti-SARS-CoV-2 antibodies with high sensitivity and specificity for each patient sample.

## MATERIALS & METHODS

### Recombinant proteins

The procedure for protein expression and production of the selected spike and RBD in this study has been detailed previously in an established and available protocol^20, 21^. Briefly, recombinant proteins from optimized DNA constructs (Addgene #166010 for Spike, Addgene #166019 for RBD) were produced in an Expi293F mammalian expression system (Thermo Fisher Scientific). After 96 hours (Spike protein) or 72 hours (RBD protein) post-transfection, supernatants from transfected cells were harvested by centrifugation, clarified, and subjected to tangential flow filtration (TFF) prior to purification using immobilized metal affinity chromatography (IMAC). Spike proteins were desalted, and RBD proteins were further purified by size exclusion chromatography. Specific details of protein production are described by Esposito et al.^20, 21^. Final proteins were analyzed and quality-checked by SDS-PAGE with Coomassie-staining, analytical size-exclusion chromatography, and mass spectrometry. Final purified proteins were aliquoted, flash frozen in liquid nitrogen, and stored at -80°C.

### Study Design & Sample Collection

Between April to October 2020, a total of 2,542 human serum samples were obtained as part of an ongoing National Highway Traffic Safety Administration (NHTSA) study of drug prevalence among adult (age 18+) trauma victims who were transported by EMS due to the severity of their injuries and had a trauma team activated/alerted at selected Level-1 trauma centers^22-24^. The specimens from this convenience sample were available for research purposes from patients who were already having blood drawn as part of medical treatment at the trauma centers. The toxicological analysis study followed the NHTSA’s standard panel for drugs known to impair psychomotor skills that could affect driving safety. When possible, excess serum samples from the study were made available for the serological analyses. Samples were collected at six study sites in the United States: Baltimore, Maryland (28.13%), Jacksonville, Florida (18.37%), Worcester, Massachusetts (10.70%), Charlotte, North Carolina (19.43%), Miami, Florida (16.48%), and Iowa City, Iowa (6.88%). The study was conducted in accordance with Good Clinical Practice, the principles of the Belmont Report and HHS regulations enumerated under 45 CFR 46. The Chesapeake/Advarra Institutional Review Board served as the central IRB for five sites, and the University of Florida Institutional Review Board served as the IRB of record for the Jacksonsville, FL site. De-identified samples and other data were included in the study under IRB-approved waivers of consent and authorization. All demographic information was obtained from medical records or other secondary sources such as emergency medical services run reports and crash reports. De-identified samples were then sent to NIH for SARS-CoV-2 ELISA testing on dry ice overnight and stored at -80°C until processing.

### Sample and control preparation

Serum samples were heated at 56 °C for 1h before use to reduce the risk from any potential residual virus in the serum. The day before running ELISA, serum samples were diluted 1:400 in blocking buffer consisting of 1xPBS + 0.05% Tween20 (PBS-T) with 5.0% Nonfat Dry Milk and can be stored in 4 °C for up to 12 hours. There were four controls in technical duplicate on each plate: blank controls used for the secondary antibody signal only, SARS-CoV-2 convalescent patient sera diluted at 1: 1000 and 1: 2500 as positive controls, and archival serum as a negative control (1:400 dilution in blocking buffer). Archival serum used as negative control were collected prior to the emergence of SARS-CoV-2.

### Enzyme-linked immunosorbent assay

The ELISA protocol was adapted from previously established protocols^18, 19, 25, 26^. This procedure utilized a semi-automated setup (BioTek Instruments EL406 washer/dispenser/stacker). High-absorption 96-well plates (NuncMaxiSorp ELISA plates; ThermoFisher) were coated with 100 µl per well of spike (1 μg/ml) or RBD (2 μg/ml) protein suspended in 1xPBS (Gibco) and incubated overnight for at least 16 hours at 4 °C. The protein solution was removed and plates were washed with 300 µL of PBS-T (0.05% Tween 20 in 1xPBS) per well three times and blocked at room temperature for 2 hours with 100 µL per well of blocking buffer (5.0% Nonfat dry milk in PBS-T). After blocking, plates were again washed three times with 300 μL of PBS-T per well. Next, 100 µl of each sample dilution or control was added in technical duplicate into the plates and incubated for 1 hour at room temperature. After sample incubation, plates were washed three times with 300 μL of PBS-T per well. Then, goat anti-human IgA, IgM, and IgG horseradish peroxidase (HRP) secondary antibodies (ThermoFisher) were diluted at 1:4000 in blocking buffer and 100 μL of each secondary antibody solution was added to each well for 1 hour. Plates were again washed three times with PBS-T, then incubated with 100 μL of 1-Step^™^ Ultra TMB-ELISA Substrate Solution (ThermoFisher) for 10 minutes followed by 100 μL of 1 N sulfuric acid STOP Solution (ThermoFisher). Within 30 minutes after adding STOP solution, optical density (OD) was measured at 450 and 650 nm using BioTek Epoch2 plate reader. To remove background, the actual absorbance was calculated as the difference between OD at 450 nm and at 650 nm before further statistical analysis.

### Statistical analysis

Seropositivity was defined as either IgG or IgM OD levels above their respective thresholds for both the spike and RBD expression. Using both spike and RBD expression together increased sensitivity and specificity to 100% for both IgG and IgM based on evaluation with convalescent positive and archival negative controls^18, 19^. The method to determine thresholds was detailed previously using simulations of different samples and control size to model the statistical confidence over a range of disease prevalence and assay specificity. The threshold was determined as previously reported to ensure that the lower 95% confidence limit of specificity is greater than 99%. Exact binomial methods were used to compare seroprevalence between population subgroups. Multiple comparisons were corrected for using the Bonferroni method. To evaluate the association between drug exposure (each drug separately, drug classes, and any drug positivity overall) and SARS-CoV-2 serostatus, multivariable penalized likelihood logistic regression was used, adjusting for age of the trauma patient, sex, race, ethnicity, emergency room admission month, and the admission city among individuals whose samples were tested for the presence of drugs and alcohol. Analysis was done using the “logistf” package version 1.24 in R version 4.0.4^27, 28^.

## RESULTS

### Cohort Characteristics

The study consists of 1434 participants identified as White (56.41%), 891 as Black or African American (35.05%), 24 as Asian or Asian American (0.94%,) 17 as Native American or Alaska Native (0.67%), 1 as Native Hawaiian or Pacific Islander (0.04%), and 175 as another race or undisclosed (6.88%). Of the study participants, four hundred and eight were identified as Hispanic or Latino (16.05%), while 2106 were identified as Not Hispanic or Latino (82.85%), and 28 unknown (1.10%). The median age was 41, with 54.64 % of participants between the ages of 18 and 44, 32.61% between the ages of 45 and 69, and 12.71% ages 70 or older. Most patients were male (72.80%) as opposed to female (26.04%). When compared to the trauma statistics generated from total admissions recorded in the trauma registry at each of the six sites during the same collection time period, the sample demographics were representative of the overall trauma population within these study sites (**Fig. 1b**). When compared to the general US population this trauma population is in general younger and contains more males and more non-white study participants, though this population is specific to the service areas within the bounds of the six trauma centers. Within the sample populations we were able to identify antibodies against the SARS-CoV-2 full spike ectodomain (spike) and spike receptor binding domain (RBD) with IgG, IgM and IgA classes (**Fig. 1c-e**). Daily measurements of seropositivity and samples collected are displayed in **Fig. 1f**, with monthly estimates in **Fig. 1g**.

**Figure 1:**
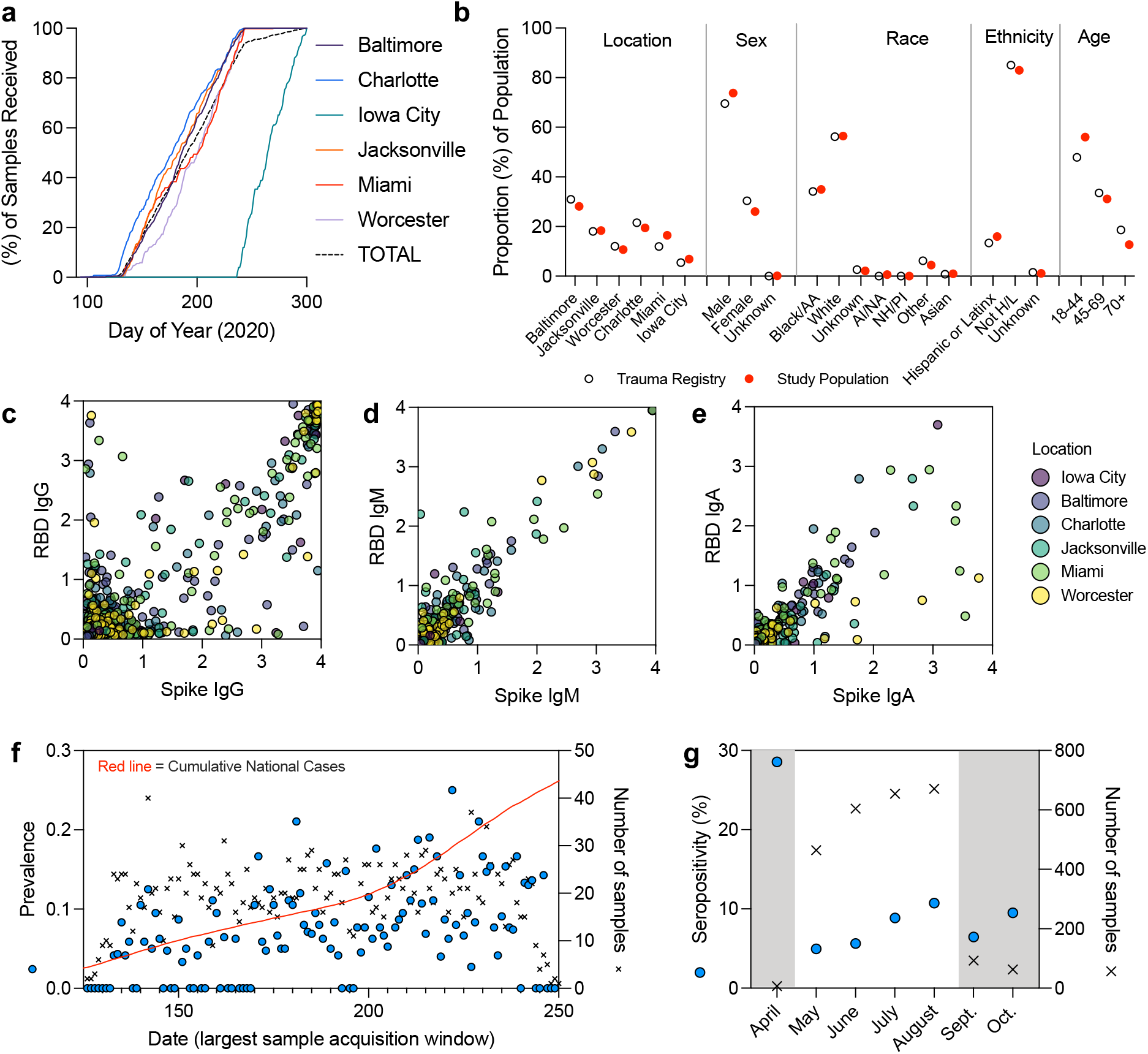
Trauma patient plasma sample collection timeline and SARS-CoV-2 serologic analysis. (a) sample collection timeline from six participating trauma centers, Baltimore (purple), Charlotte (blue), Iowa City (green), Jacksonville (red), Miami (orange), Worcester (light purple), total (dashed black). (b) Comparison of study population (red dot) to trauma registry data (open circle) within the same timeframe of collection. (c) Raw IgG serology ELISA absorbance values for SARS-CoV-2 Spike ectodomain, and receptor binding domain (RBD), (d) IgM, (e) IgA. (f) Number of samples collected (black x) versus daily seroprevalence (blue circle, see statistical methods) in the context of overall US national case trends (red line) during the main collection window. (g) Monthly seropositivity of samples, main collection window in white.

### Anti-SARS-CoV-2 isotype profile among seropositive participants

A range of different antibody isotype profiles were detected against both full spike ectodomain (spike, **Fig. 2a**) and spike receptor binding domain (RBD, **Fig. 2b**) antigens. A positive correlation (p ≥ 0.000001) was found with all isotypes tested, with the strongest correlations between IgG and IgA isotypes and lower correlation of IgG with IgM (**Fig. 2c**). The majority of those who tested positive were IgG positive (IgG+, **Fig 2. d-f**, n = 188/226 or 83.19%). Of those that were IgG+, more than half (51.60%) had high concentrations of antibody in their serum as measured by an OD reading greater than three, which correlates with a monoclonal recombinant anti-RBD human IgG antibody concentration of > 150 ug/ml^17^. IgM and IgA overall had lower concentrations of antibody in comparison to IgG, in agreement with previous findings in the literature.

**Figure 2:**
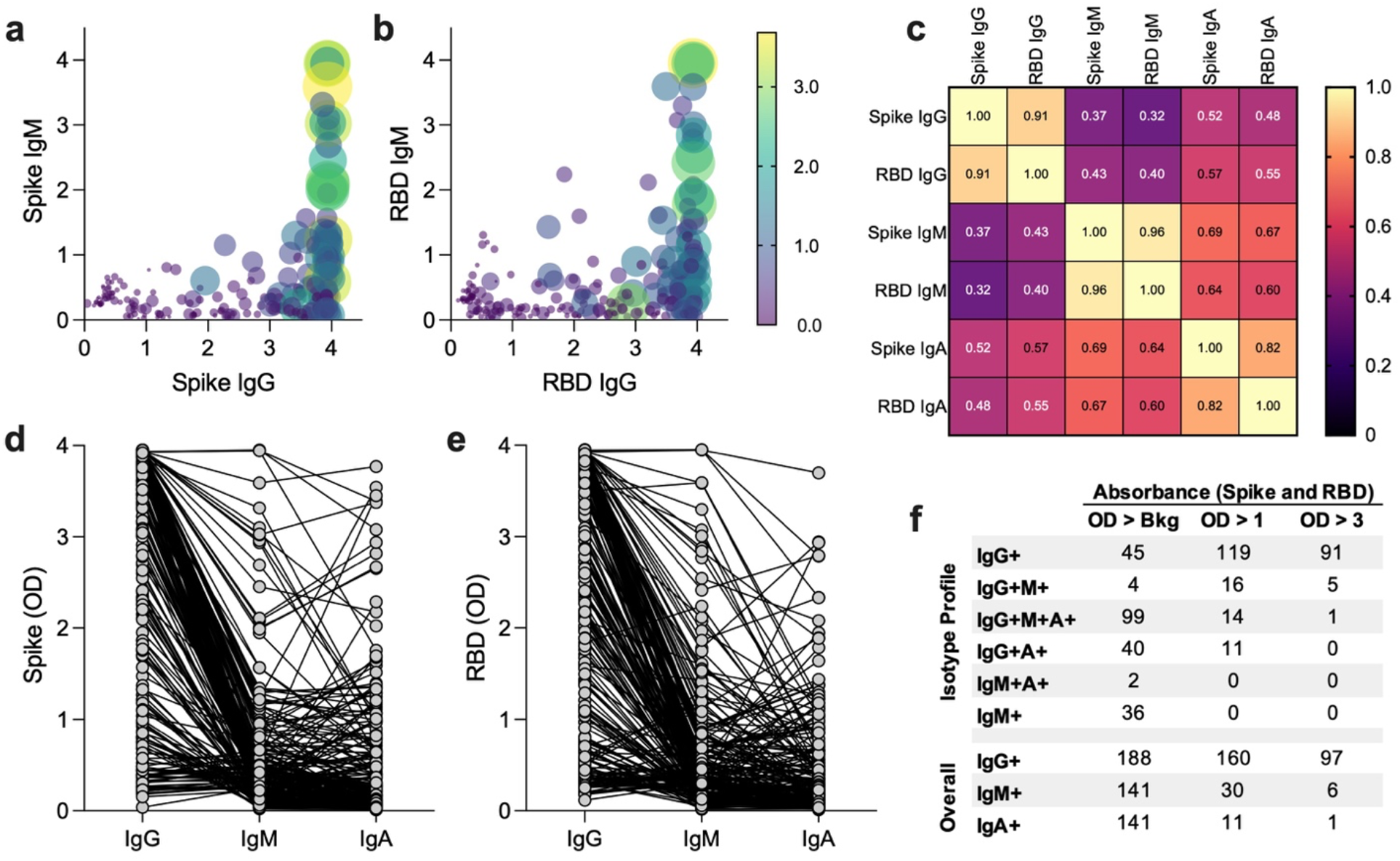
SARS-CoV-2 antibody isotype profile in seropositive patients. (a) Absorbance values for spike IgG (x-axis), IgM (y-axis), and IgA (point size/color). (b) Absorbance values for RBD IgG (x-axis), IgM (y-axis), and IgA (point size/color). (c) Correlation of expression between different serologic analytes. (d-e) Individual antibody comparing OD levels of IgG, IgM, and IgA isotypes for (d) spike and (e) RBD analytes. (f) Intensity of ELISA reading with “Bkg” = Threshold/Background, OD >1 being mid to high antibody concentration, and OD > 3 representing high and off-scale high antibody concentrations.

### SARS-CoV-2 Seroprevalence in Trauma Patients from Six Sites by Demographic Groupings

Overall, 7.87% (95% CI: 6.88 – 8.98) of participants were seropositive (**Fig. 3**). The highest seroprevalence point estimate was in Miami (12.17%, 9.36 – 15.67). Male and female participants had similar seroprevalence (male: 7.73%, 6.60 – 9.03; female: 8.31%, 6.42 – 10.67). Black/African American participants had the highest seropositivity of any race (9.54%, 7.77 – 11.65) which is significantly higher than the overall estimate (Bonferroni adjusted *p* <0.05), and significantly higher (p = 0.0001) than White participants (5.72%, 4.62 – 7.05). In addition, Hispanic/Latino participants (14.95%, 11.8 – 18.75) had the highest seroprevalence point estimate of any demographic group, which was significantly higher (p = 0.0001) than non-Hispanic/Latino participants (6.55%, 5.57 – 7.69). The youngest age group, 18 – 44 years, had significantly higher seroprevalence (9.79%, 8.33 – 11.47) than both older groups: 45 – 69 years (6.03%, 4.59 – 7.88, p = 0.0004) and 70+ years (4.33%, 2.54 – 7.20, p = 0.0051), respectively.

**Figure 3:**
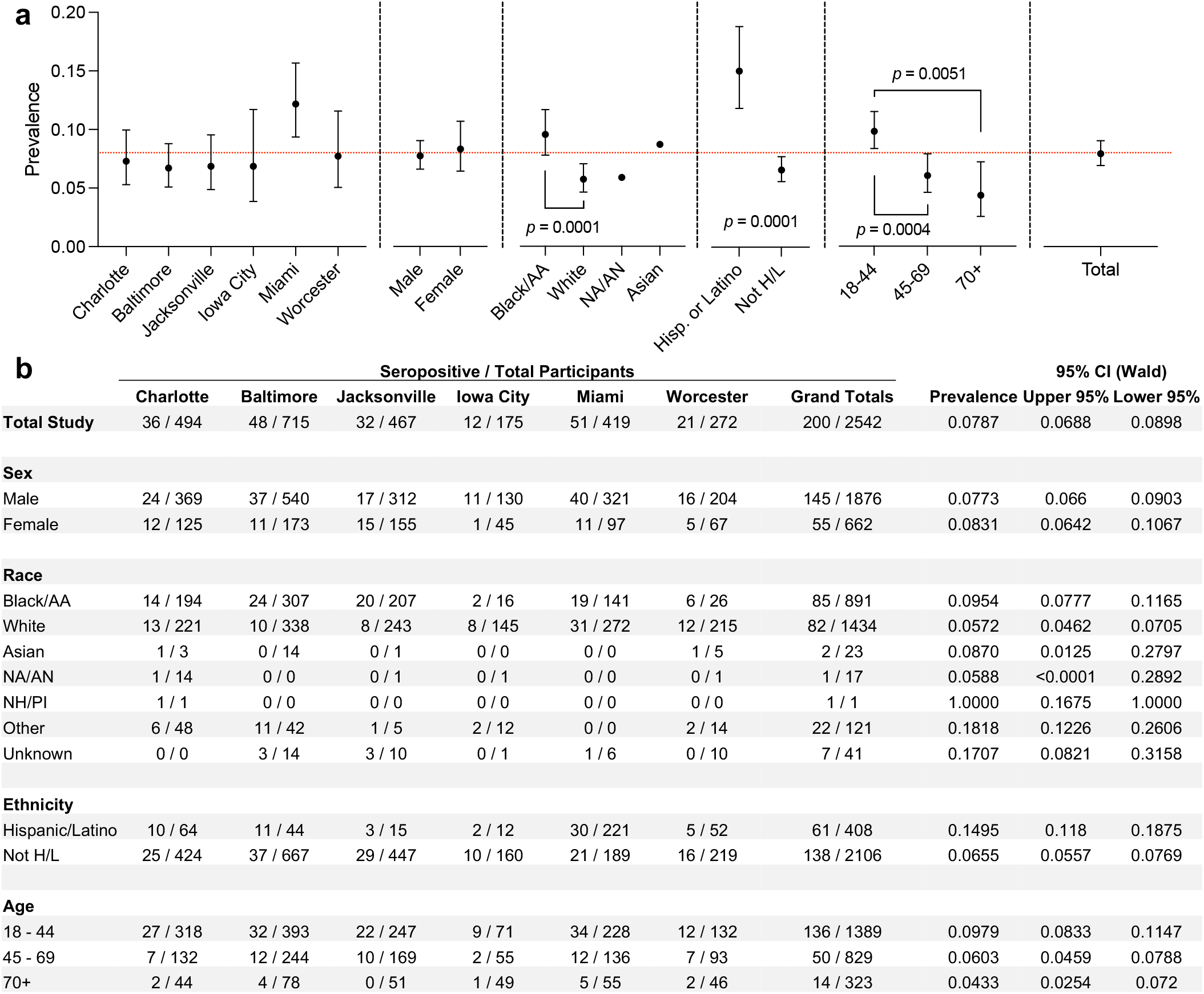
Seroprevalence of SARS-CoV-2 in Trauma Patients During the Summer 2020 COVID-19 wave. (a) Seroprevalence of SARS-CoV-2 antibodies by demographics, data are means ± 95% confidence interval (Wald), CI’s not shown for large-error (small n) samples (NA/AN, Asian). Significance = students T-test, determined by Bonferroni post-hoc adjustment for multiple comparisons. Red line = total seroprevalence of overall study population. (b) Chart of raw seropositivity data from different demographic groupings within different sites. AA = African American; NA/AN = Native American/Alaska Native; H/L = Hispanic or Latino; NH/PI = Native Hawaiian/Pacific Islander.

Of these participants, 1,679 were tested by PCR for active SARS-CoV-2 infection on site at the trauma centers. Testing approaches and rates varied by site due to differences in the availability of testing materials at the trauma centers. Of those tested for active infections, 71 patients (4.23%, 3.36 – 5.31) were positive for SARS-CoV-2. Within this group of 71 identified active infections, 41 cases were seropositive (seropositivity of participants with positive COVID test: 57.75%, 46.14 – 68.55), and the other 30 cases were seronegative (42.25%). This suggests that the majority of participants that tested positive for COVID were convalescent and most likely outside of the window of when they were most infectious; however, over 40% of these participants were pre-convalescent (seronegative) suggesting early stages of disease which is associated with higher viral loads.

### Trauma characteristics and correlations with seropositivity

The highest number of admitted trauma patients were motor vehicle crash victims (MVC’s; *n* = 1162, 46.18 %) followed by falls (*n* = 601, 23.89 %), firearm injury (GSW; *n* = 273, 10.85 %), stab (*n* = 91, 3.62 %), assault (*n* = 91, 3.62 %), drowning (*n* = 1, 0.04 %), fire/burn (*n* = 30, 1.19 %) and other traumas (*n* = 190, 7.55 %) (**Fig. 4**). When calculating seroprevalence point estimates, “other motorized transport injuries” were grouped with MVC’s as road traffic injuries (RTI), while drowning and fire/burn injuries were categorized as “other” (**Fig. 4a**). Assaults had the highest seroprevalence at 14.28% (8.4 – 23.06), though this was not significantly higher than for other trauma categories. RTIs, falls, and other traumas had similar seroprevalence (MC: 7.7%, 6.33 – 9.36; fall: 6.49%, 4.76 – 8.77; other: 6.78%, 4.08 – 10.97), while seroprevalence among firearm injuries and stab wounds was slightly higher (GSW: 9.89%, 6.84 – 14.05; stab: 9.68%, 5.49 – 16.29). Overall, type of trauma was not significantly associated with seroprevalence.

**Figure 4:**
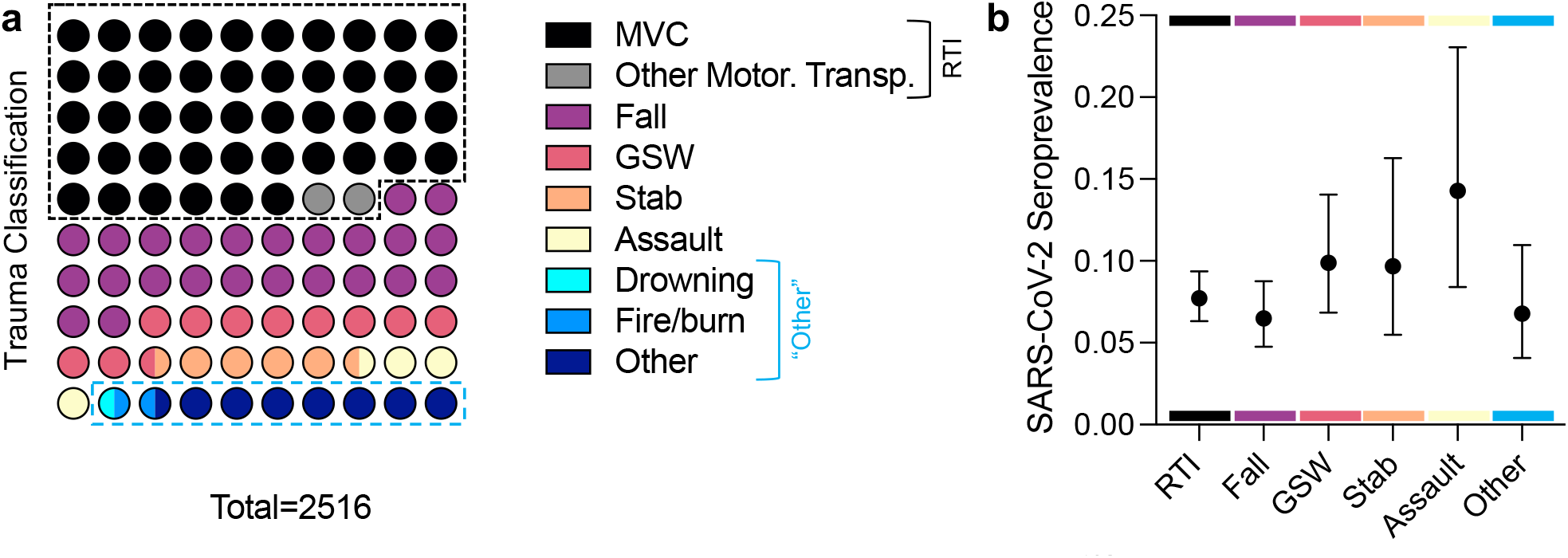
Trauma classifications and correlations with SARS-CoV-2 seropositivity. (a) Trauma classification of 2516 trauma cases. (b) SARS-CoV-2 seroprevalence in different trauma groups. Road Traffic Injuries (RTI) = Motor Vehicle crash (MVC) and Other Motorized Transportation. GSW = Gunshot Wound. Other = classified as “other” plus Drowning and Fire/Burn cases (due to low *n*). Data are point estimates ± 95% confidence interval (Wald).

### Correlation of drug use with prior SARS-CoV-2 infection

Drug toxicology results, based on the NHTSA’s standard screening for drugs known to affect driving safety, were available for 1,162 of the motor vehicle crash victims included in the current seroprevalence study. Of these, 55.54 % tested positive for one or more drugs (52.69 – 58.37), including legal and decriminalized compounds such as alcohol and marijuana. The full list of drugs that were tested are available in **Table 1**. For further analysis, these drugs were classified in larger groupings as stimulants, narcotics or sedatives, anti-depressants, and others classification of drugs (**Fig. 5)**.

**Table 1:**
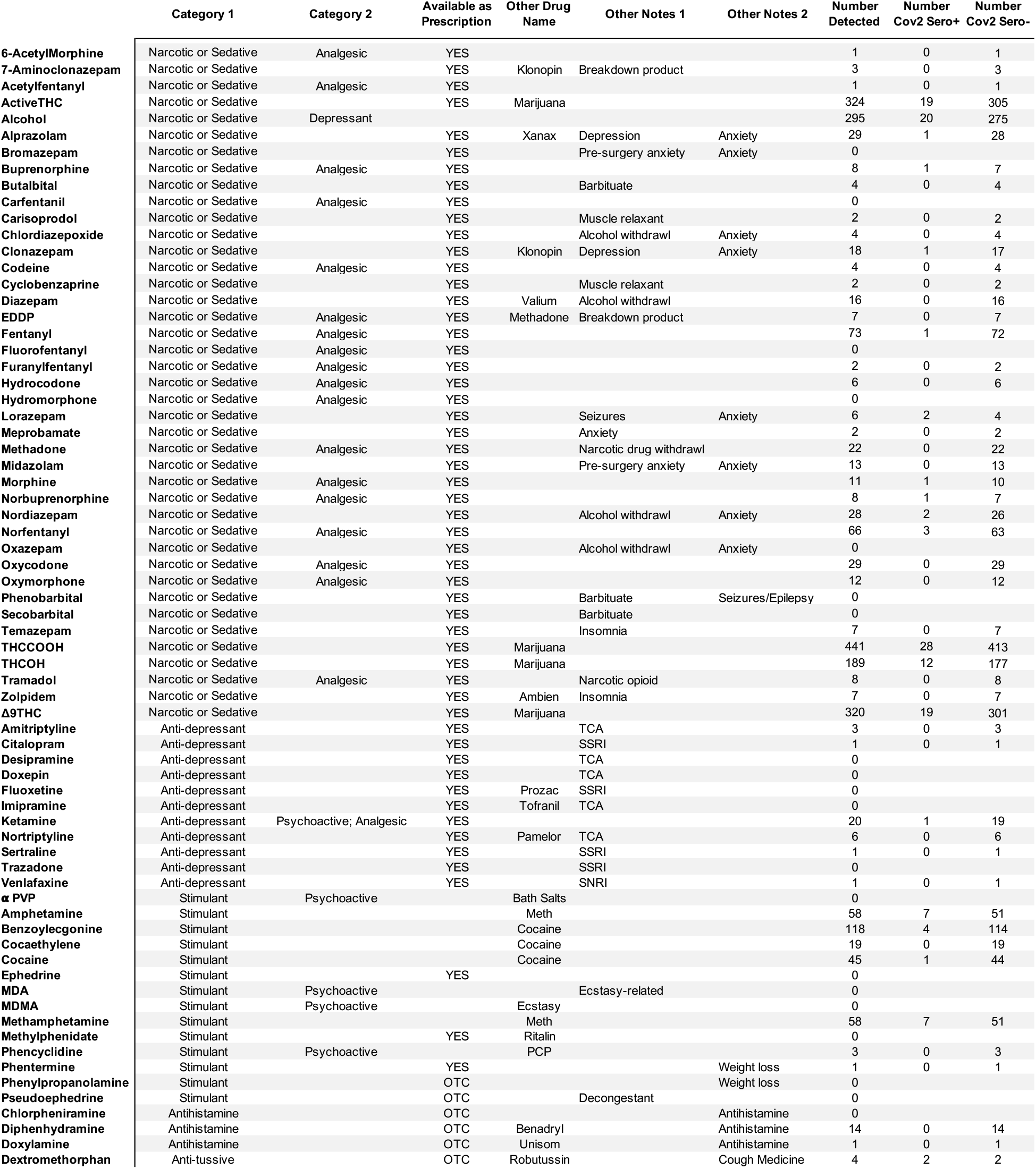
Drugs evaluated in trauma patients and seropositivity results. Details regarding the individual drugs are displayed along with other drug name and notes regarding its application. The number detected is the number of samples that tested positive for the given drug. Number Cov2 Sero+ is the number that tested positive for IgG or IgM antibodies against SARS-CoV-2 spike protein. OTC = over the counter.

**Figure 5:**
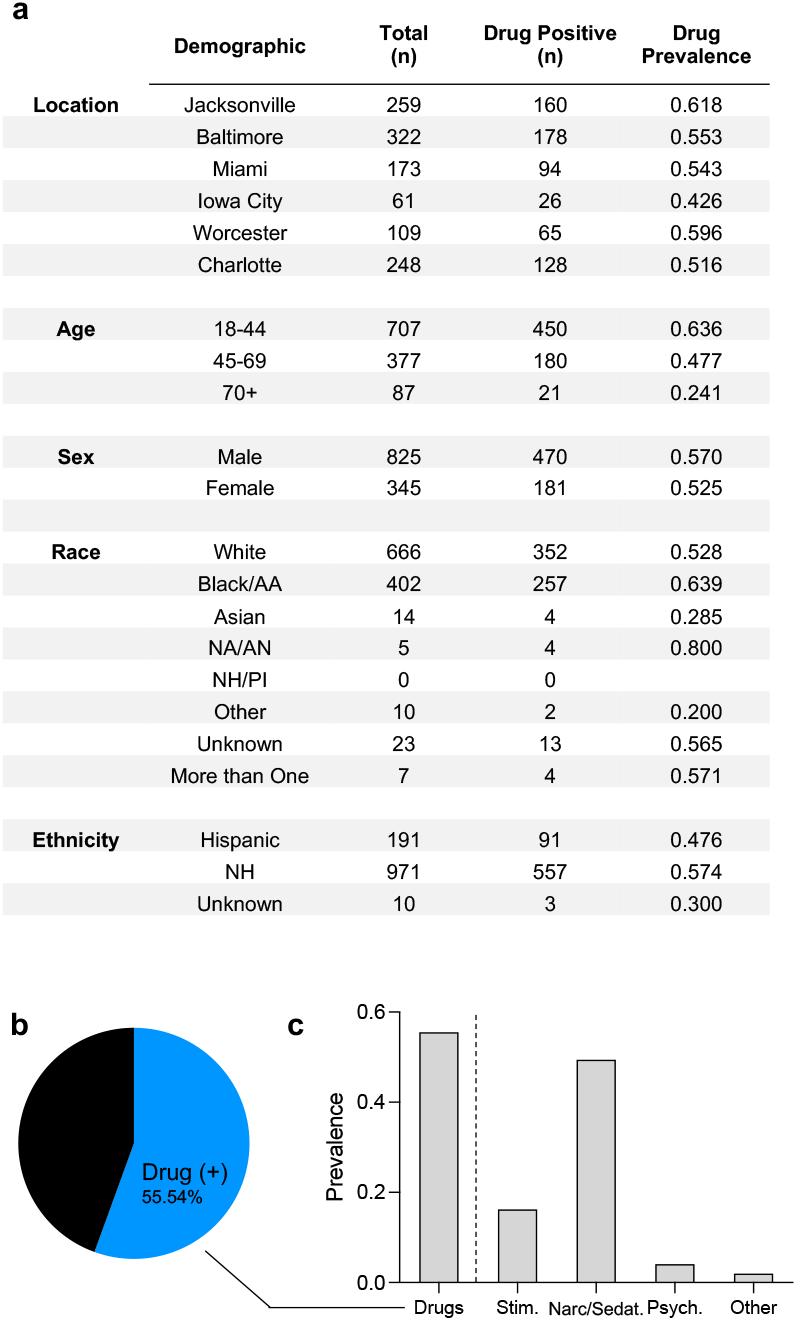
Drug prevalence in trauma patients during the summer 2020 COVID-19 wave. (a) Chart of number of trauma cases tested for drugs (motor vehicle crashes only). (b) Proportion of trauma victims that tested positive for one or more drugs in blood plasma. (c) Prevalence of different drug classifications within the trauma population^14^. Stim. = stimulants; Narc/Sedat. = Narcotic or Sedatives; Psych = Psychoactive.

Only two of the individual drugs tested were significantly associated with SARS-CoV-2 seropositivity after controlling for potential confounders. Samples tested positive for Lorazepam – belongs to a class of drugs known as benzodiazepines – were associated with an increased likelihood of being SARS-CoV-2 seropositive (Odds Ratio (OR): 8.14, 95% CI: 1.21 – 45.0, p = 0.03). Meanwhile, samples tested positive for fentanyl, a synthetic opioid class, were associated with a decreased likelihood of being SARS-CoV-2 seropositive (OR: 0.25, 95% CI: 0.03 – 0.95, p = 0.04). Narcotics or sedatives as a category were also negatively associated with SARS-CoV-2 seropositivity (OR: 0.56, 95% CI: 0.34 – 0.90) (**Fig. 6**). When comparing drug positive versus drug negative patients (those that overall had drugs detected in their toxicology), there was a slight positive trend between anti-depressant positivity and SARS-CoV-2 seropositivity, though there was no significant difference. However, samples that tested positive for narcotics or sedatives had a significantly negative correlation with SARS-CoV-2 seropositivity (p = 0.018).

**Figure 6:**
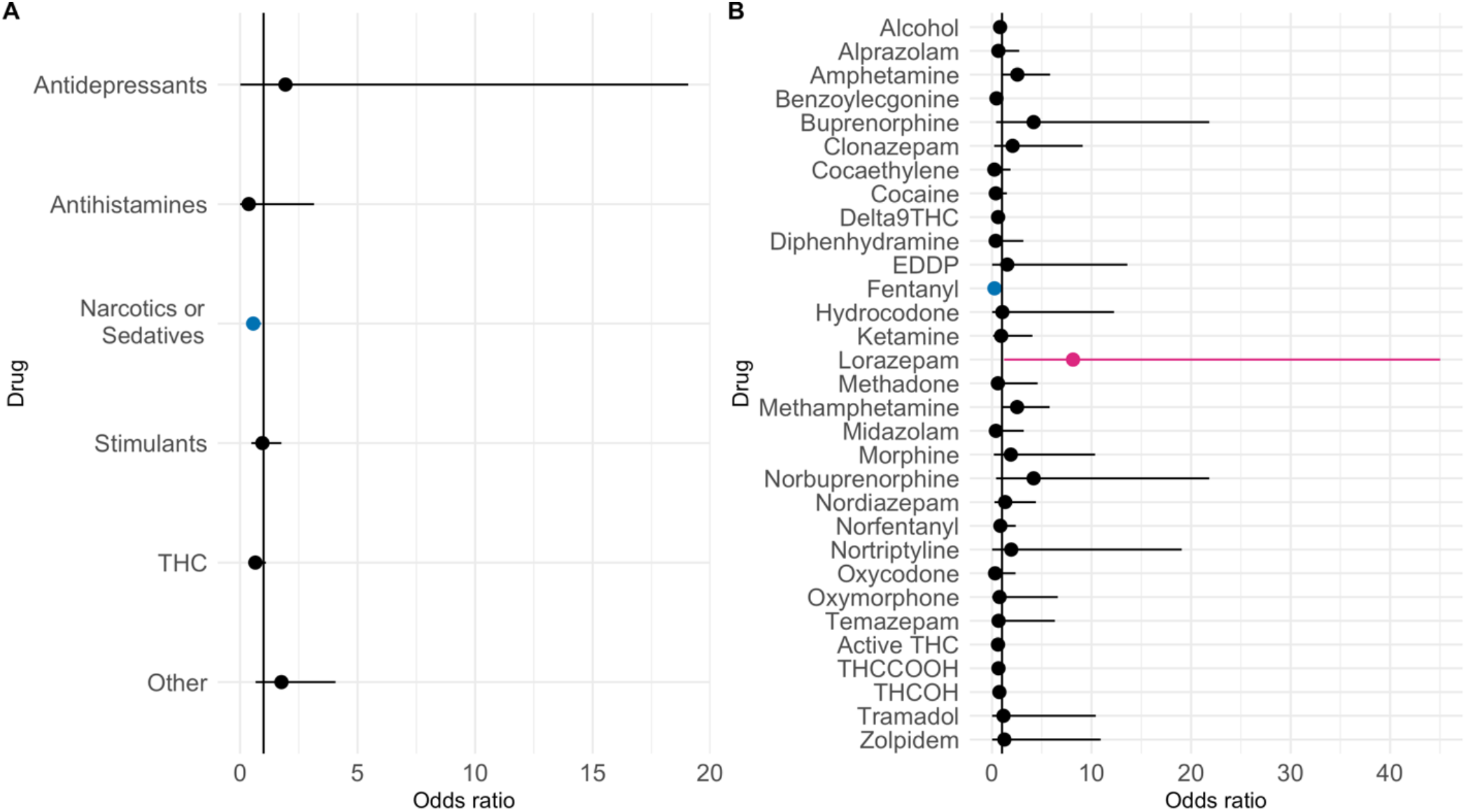
Correlation of SARS-CoV-2 Seroprevalence and Drug Presence in Trauma Patients. Odds ratios with 95% confidence intervals of a positive drug test and a positive SARS-CoV-2 seropositivity result. (a) Drug Classes. (b) Individual drugs. Red = positive correlation. Blue = Negative correlation. Black = not statistically significant.

## DISCUSSION

Exposure of healthcare workers and first responders to infectious diseases can be concerning for both their own health and safety, how they ensure proper care for an infected patient, as well as the health and safety of other patients. During the SARS-CoV-2 pandemic, we have witnessed the personal protective equipment shortages that can put both providers and patients at risk. One instance where control and isolation in the context of potential infectious diseases is difficult to maintain is in trauma where the main goal is to stabilize a patient’s potentially life-threatening injuries^29^. Trauma itself could also have a negative effect on the patient’s immune response against an infectious pathogen due to long-term immune dysfunction associated with traumatic injury^30^. The emergence of a novel virus such as SARS-CoV-2 gives the research community an opportunity to thoroughly characterize the potential differential burden of respiratory viruses in trauma patients at the time of admission. As such, we evaluated the prevalence of SARS-CoV-2 in trauma patients both for knowledge regarding the COVID-19 pandemic as well as data to inform the medical field of potential considerations for other respiratory viruses. In our study population of patients from urban U.S. trauma centers, we found similar and in some cases exaggerated seroprevalence compared to other seroprevalence studies in the United States^18, 31-40^. In this cohort, trauma patients who were SARS-CoV-2 seropositive were more frequently Hispanic or Black/African American, and young (< 45 years). Over 45% of seropositive trauma patients under the age of 45 identified as Black/African American, and 33% were Hispanic/Latino.

While it is difficult to compare the relative risk of SARS-CoV-2 seropositivity between the general population and the trauma population due to differences in donor recruitment and study design, we did note that in comparison to a national study conducted by our group using the same seroassays over the same time period, cities in the south/central region of the United States had higher SARS-CoV-2 prevalence in trauma patients comparison to the general population. As the samples represented this trauma population were obtained from only six trauma centers and not all Level-1 trauma facilities throughout the United States, the study cannot be used to infer seroprevalence in the overall trauma population of the United States. In addition, one of six sites, Iowa City, had a slightly delayed collection timeframe in comparison to the other five. Therefore, further investigation is necessary to understand the seroprevalence at each trauma center in comparison to its region.

Published seroprevalence estimates of the general population of Massachusetts during the same time period as this study showed a lower prevalence (4.0%) in comparison to the trauma population sampled from Worcester, MA (7.72%)^39^. Another study in Miami found that during the spring/summer of 2020 the general population had a lower seroprevalence of 6.0% compared to the trauma population at 12.17%^41^. A study in central North Carolina found increasing seroprevalence in the general community from 2.9% to 9.1% from April through October. Our estimate from Charlotte, NC which was gathered in July at 7.28%, could suggest a higher than state-average rate of SARS-CoV-2 seroprevalence in the trauma population, given the steady rate of new case diagnosis in North Carolina during this timeframe, though further analyses are needed to evaluate the probability of this phenomenon^5^.

Among motor vehicle crash victims specifically, a large proportion of trauma victims were positive for drugs or alcohol. There was a lower likelihood that these individuals had a prior SARS-CoV-2 infection if they were positive for narcotics or sedatives (including marijuana). Interestingly, there was a positive correlation with the depressant Lorazepam and SARS-CoV-2 seropositivity; this medication induces anxiolysis and sedation. In addition lorazepam can worsen obstructive pulmonary disease and lead to respiratory compromise^42^. Whether seropositivity among specific drug exposed individuals is due to chemical activity of the drug or alterations in behavior of those using these drugs remains to be determined, as do the implications of these relationships for first responders and other medical professionals needing to treat trauma patients under the influence of certain drugs. As the stimulant drug class trends higher seropositivity than THC, alcohol, benzodiazepines, or narcotics, and methamphetamine use is correlated with increased risk-taking behavior, this may explain a higher trending SARS-CoV-2 exposure. Additionally, patients using stimulants such as methamphetamine often present in the emergency room with excited delirium, spitting, and physical aggression can lead to breakdown in PPE protocols for healthcare providers.

The increased incidence of previously reported viral infections (HIV and hepatitis) in trauma victims could be co-dependent upon the increased prevalence of injectable drug use in the trauma population, creating difficulty in determining correlation versus causation^43, 44^. Given that SARS-CoV-2 is a respiratory pathogen and as of the writing of this manuscript not known to be transmitted by blood or needle sharing, this could create an important consideration for other respiratory viruses, such as influenza, necessitating an evaluation of personal protective equipment afforded to first responders, and considerations in patient care for trauma patients.

In this study, we have shown that differences in SARS-CoV-2 seroprevalence among trauma patients are, as with the general population, correlated with region, race, ethnicity, and age. There are also correlations associated with use of legal and illegal drugs, including a negative correlation of SARS-CoV-2 seropositivity with the use of narcotics or sedatives. A number of factors can affect respiratory disease spread and severity from the population level to the individual level. More densley populated areas can be subject to more rapid spread of disease due to increased likelihood of coming into contact with an infected individual^45, 46^. Unequal access to healthcare and education on disease prevention can also lead to differences in disease spread^47^. Prior to the COVID-19 outbreak, trauma patients have been shown to have higher incidence of a variety of infectious diseases, though prior research has focused on bloodborne pathogens. Respiratory diseases such as SARS-CoV-2 have the potential to complicate care plans for trauma patients who are susceptible to increased risk for post-trauma lung conditions such as pneumonia. Our results suggest a potential higher incidence of SARS-CoV-2 in trauma patients. These data are important in evaluating both the varying risks that are posed to first responders, as well as understanding potential patterns of infectious disease spread to prepare seasonal and future emerging infectious disease threats in at-risk patients.

## Data Availability

Data are available upon reasonable request to the corresponding author after article publication.

## ACKNOWLEDGEMENTS

The authors would like to acknowledge Dr. Corbett and Dr. Graham of the NIAID VRC for their generous donation of coronavirus spike expression plasmid, and Dr. Aaron Schmidt, J Feldman, BM Hauser and T M Caradonna of the Ragon Institute of MGH, MIT, and Harvard for their donation of their RBD expression plasmid.

## Funding

This research was supported in part by the Intramural Research Program of the NIH, including the National Institute for Biomedical Imaging and Bioengineering, the National Institute of Allergy and Infectious Disease, and the National Center for Advancing Translational Sciences. This project has been funded in part with Federal funds from the National Cancer Institute, National Institutes of Health, under contract number HHSN261200800001E. The content of this publication does not necessarily reflect the views or policies of the Department of Health and Human Services, nor does mention of trade names, commercial products, or organizations imply endorsement by the U.S. Government.

### Disclaimer

The NIH, its officers, and employees do not recommend or endorse any company, product, or service.

